# Long-term prognosis of patellofemoral pain in adolescents and adults: A systematic review with meta-analysis and meta-regression

**DOI:** 10.64898/2026.04.27.26351023

**Authors:** KD Lyng, EM Machado, MB Blumenfeld, S Gürühan, A Andreucci, LB Sørensen, N Pourbordbari, C Vad, CL Straszek, SK Johansen, MS Rathleff, G Vasconcelos

**Affiliations:** Department of Health Science and Technology, Faculty of Medicine, Aalborg University, Denmark; Center for General Practice at Aalborg University, Department of Clinical Medicine, Aalborg University; Department of Physical Therapy, Federal University of São Carlos, São Carlos, Brazil; Bitlis Eren University, Faculty of Health Sciences, Department of Physiotherapy and Rehabilitation, Bitlis, Turkey; Department of Physiotherapy, University College of Northern Denmark (UCN), Aalborg, Denmark; Department of Functional Health, Federal University of Goias, Goiânia, Brazil

## Abstract

**Objective:** To investigate the long-term (defined as ≥12 months) prognosis of knee pain and knee function in adults and adolescents with patellofemoral pain (PFP).

**Design:** Systematic review with meta-analysis and meta-regressions.

**Data sources:** MEDLINE, OVID, CENTRAL, Web of Science, OpenGrey, and International Patellofemoral Research Retreat abstract books.

**Eligibility criteria for selecting studies:** Prospective studies of patients clinically diagnosed with PFP, aged <40 years, with a long-term follow-up (minimum of 12 months). Primary outcomes were self-reported pain intensity (worst, during activity, and usual) and function. Meta-analyses and meta-regressions were performed where appropriate. Narrative synthesis was performed for those not included in the metanalysis. Risk of bias was assessed using the Quality In Prognosis Studies (QUIPS) tool, and certainty of evidence using GRADE.

**Results:** A total of 42 studies (n = 3,230) were included. At 12 months, meta-analysis indicated reduction in worst pain (SMD 1.36; 95% CI 0.85–1.86), pain during activity (SMD 1.36; 95% CI 0.61–2.11), and resting pain (SMD 0.91; 95% CI: 0.75- 1.08). No significant reduction was found for usual pain. We found improvement in self-reported function (investigated using the Anterior Knee Pain Scale (AKPS) MD 14.60; 95% CI 11.60–17.61), FIQ (MD 3.33; 95% CI: 2.46- 4.20) and the Western Ontario McMaster Universities Osteoarthritis Index (WOMAC) (MD –7.73; 95% CI: -10.36 to – 5.10). Extended follow-up (≥60 months) suggested more variable improvements. Meta-regression showed no association between age and 12-month function, while older age was modestly associated with greater improvement in activity-related pain at extended follow-up. Overall, a considerable proportion of participants continued to report persistent symptoms, and heterogeneity across studies was substantial. Certainty of evidence ranged from very low to moderate across outcomes investigated.

**Conclusion:** Pain and self-reported function generally improve over time, particularly within the first 12 months. However, substantial heterogeneity and persistent symptoms in a considerable proportion of patients at extended follow-up indicate that recovery is not universal and trajectories are highly variable.

**What is already known:**

- Patellofemoral Pain (PFP) is a very common condition in both adolescents and adults.
- Multiple treatments modalities exist, including patient education and exercise therapy.
- People suffering from PFP request more knowledge on the long-term prognosis.

**What are the new findings?:**

- This systematic review and meta-analysis provide the most comprehensive synthesis to date of long-term outcomes (≥12 months) in adolescents and adults with patellofemoral pain.
- Pain and self-reported knee function generally improve at the group level over time, particularly within the first 12 months.
- Despite group-level improvement, a substantial proportion of individuals continue to report persistent symptoms, indicating that patellofemoral pain is often not fully self-limiting.
- Long-term outcomes are highly heterogeneous, with different pain constructs demonstrating distinct trajectories across follow-up periods.
- Meta-regression identified no consistent prognostic associations, suggesting that current study-level variables explain little of the variability in long-term outcomes.

**How might this study affect research, practice or policy?:**

- Clinicians should communicate that while improvement is common in patellofemoral pain, persistent symptoms are frequent, highlighting the need for realistic prognostic expectations and long-term management strategies.
- Future research should prioritise harmonised outcome measures and long-term follow-up to better understand recovery trajectories and identify subgroups at risk of persistent symptoms.

## INTRODUCTION

Patellofemoral pain (PFP) is one of the most common knee conditions in both adolescents and adults aged ≤ 40 years.^1^ People with PFP typically experience anterior knee pain around or behind the patella, particularly during weightbearing activities such as running, stair ambulation, or squatting.^2^ Clinically, PFP is often considered challenging as the underlying aetiology is unknown and individuals often report reduced function, impaired quality of life, and elevated psychological distress, including pain catastrophising, fear of movement, and depressive symptoms.^3–7^

A systematic review and network meta-analysis by Winters et al. supports a range of potentially relevant options, including patient education, exercise therapy, and orthoses, which have been shown to be superior to wait-and-see after 12 months.^8^ However, evidence highlights that irrespective of the treatment options used, many patients continue to report pain and symptoms even years after the initial evidence-based treatment is given.^6,9–12^ In this regard, one study showed that 40% will continue to experience pain after 12 months,^6^ another study reported that 51% will still report pain after 12 months,^12^ and a third study showed that 57% will continue to experience unfavourable outcomes after 5-8 years.^13^ Our recent patient-public engagement study highlighted that one of the top research priorities for people suffering from PFP was either “*When will my pain go away*” and “*when will I be back playing sport*”.^14^

A recent evidence and gap map by Neal et al. 2025 systematically charted the existing literature on prognostic factors in PFP and highlighted significant heterogeneity in populations, timepoints, and outcomes.^16^ Their findings underscored the need for research that focuses specifically on long-term prognosis, including pain and function outcomes that reflect patient priorities.^14^ Moreover, recent findings suggest that clinicians initial prognostic assessments of PFP are not associated with patients’ actual outcomes following exercise therapy, further highlighting the difficulty of accurately predicting individual trajectories.^15^

Currently, there is no up-to-date synthesis of the long-term prognosis of PFP, making it difficult to provide patients with a credible answer to the common question *“When will my pain go away?”*. ^14^ This highlights the need for this systematic review on the long-term trajectory of PFP. Therefore, the aim of this systematic review is to estimate the long-term prognosis of PFP, defined as knee pain and/or self-reported knee function at follow-up ≥12 months, in adolescents and adults, through quantitative synthesis using meta-analysis and meta-regression.

## METHODS

### Protocol and registration

This systematic review was conducted according to a pre-registered protocol on the Open Science Framework (DOI: 10.17605/OSF.IO/WD4T3). The review adhered to the Preferred Reporting Items for Systematic reviews and Meta-Analyses (PRISMA) Statement,^10^ the Reporting of Quantitative Patellofemoral Pain (REPORT-PFP),^11^ and the PERSiST (implementing Prisma in Exercise, Rehabilitation, Sport medicine and SporTs science) guidance.^12^

### Eligibility criteria

We included prospective studies of adolescents and adults (<40 years) with a clinical diagnosis of PFP. Eligible designs included prospective cohort studies, randomised and non-randomised controlled trials, and case series with ≥20 participants. Studies were required to report at least one long-term outcome (≥12 months) related to knee pain or self-reported function. We included both published and unpublished studies, and no date restrictions were applied. Exclusion criteria included retrospective designs, case reports or small case series (<20 participants), studies not reporting long-term outcomes (≥12 months), and non-original research (e.g., editorials, commentaries). Only English-language articles were included due to resource constraints.

### Information sources

A systematic literature search was adapted from Winters et al.^8^ for this review in collaboration with a research librarian. The search strategy included a combination of medical subject headings (MeSH) and free-text terms related to PFP and prognosis. The following databases were searched: MEDLINE via PubMed, EMBASE via OVID, Cochrane Central Register of Controlled Trials (CENTRAL), and Web of Science. To capture grey literature, we searched OpenGrey.eu and manually screened conference proceedings from the International Patellofemoral Research Retreat (2009, 2011, 2013, 2015, and 2017). The first literature search was conducted on the 22^nd^ of February 2021 and updated the 20^th^ of January 2026. The search strategy can be seen in online **Supplementary Appendix 1**.

### Source of evidence selection

All identified records were imported into EndNote (Clarivate, Philadelphia, PA, USA) for deduplication and then uploaded to Covidence (Veritas Health Innovation, Melbourne, Australia) for title/abstract and full-text screening. Title and abstract screening were conducted independently by at least two reviewers (KDL, NG, AA, LBS, CaV, MBB, SG, GV, and EM). Full texts were assessed for eligibility by the same reviewers, working in pairs. Disagreements were resolved through consensus or consultation with a senior author (MSR). Studies excluded at the full-text stage were documented with reasons in a PRISMA flow diagram (See **Figure 1**).

**FIGURE 1.**
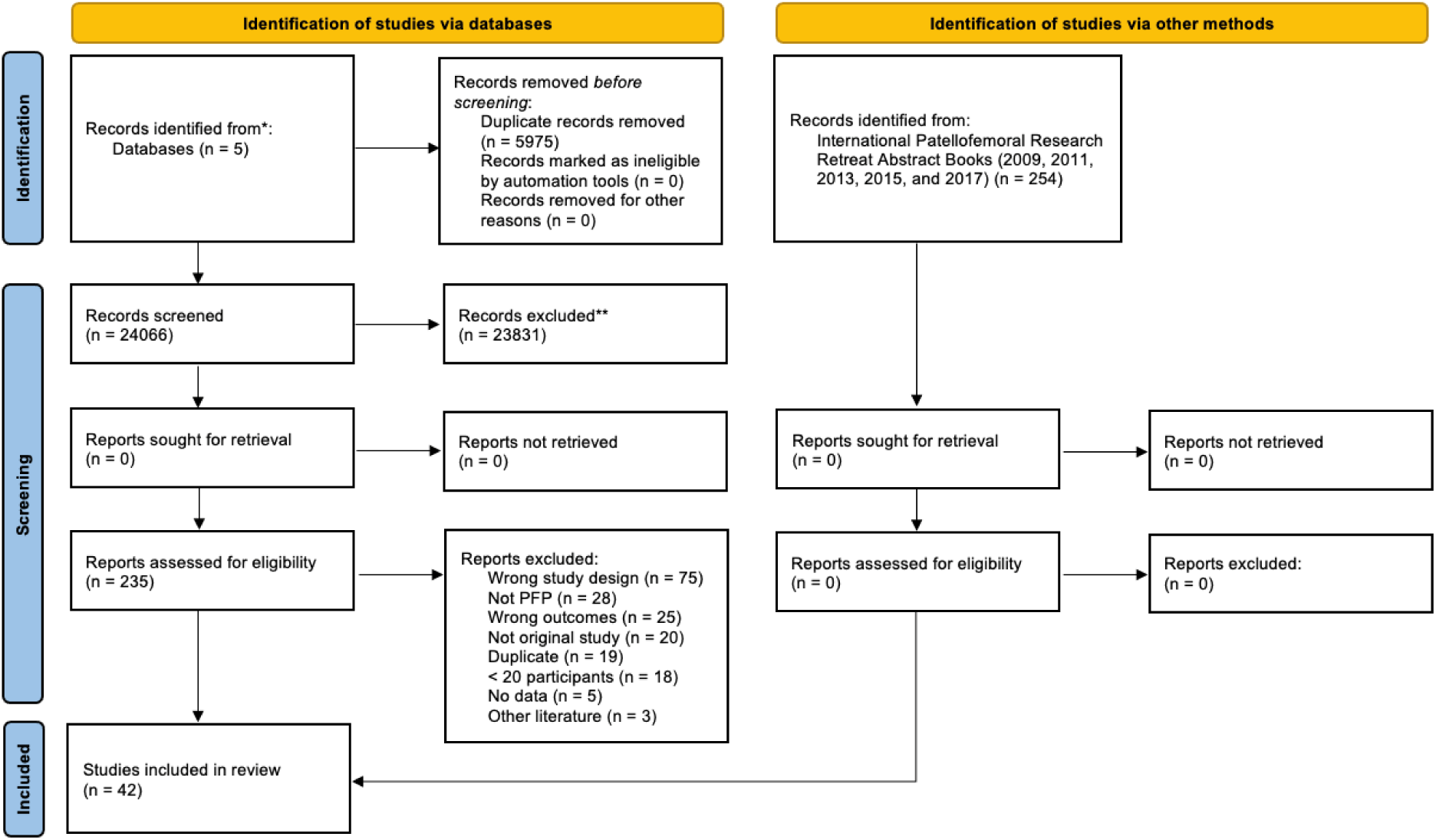
LEGEND: PRISMA FLOW CHART

### Data Extraction

Data were extracted independently by pairs of reviewers (KDL, NG, AA, LBS, CaV, MBB, SG, GV, EM, CLS, SKJ) using a pre-piloted extraction form in Microsoft Excel (Microsoft, Redmond, WA, USA). Discrepancies were resolved by discussion or adjudication by a third reviewer (MSR). Extracted information included:

- **Study characteristics:** title, main author, publication year, journal, country, design, setting, duration of the study, sample size, study design and source of financial support.
- **Participant baseline characteristics:** sex, age, BMI, symptom duration, pain intensity, pain characteristics, activity level.
- **Intervention and comparator characteristics:** type, duration, and delivery mode.
- **Follow-up:** duration, timepoints assessed, and follow-up rates.
- **Outcomes:** all reported measures related to pain (e.g., VAS (Visual Analogue Scale), NPRS (Numeric Pain Rating Scale), Function (e.g. Anterior Knee Pain Scale (AKPS), Knee injury and Osteoarthritis Outcome Score (KOOS)), Quality of life (e.g. EuroQol 5-Dimension questionnaire (EQ-5D)), Psychological factors (e.g. kinesiophobia, anxiety, depression, pain catastrophising), Self-reported recovery (e.g. Global Rating of Change (GROC)), and/or Return to sport/work and use of analgesics.

Outcomes were extracted at predefined timepoints: 12 months (±3 months), 24 months (±3 months), 36 months, and ≥60 months. We prioritised all outcomes, although the primary outcome of interest was knee pain and self-report knee function evaluated using patient-reported outcome measures. Pain outcomes were extracted and prioritised using the following predefined hierarchy: 1) worst pain (e.g. highest reported score on VAS/NPRS), 2) pain during activity (e.g. squatting, stairs), 3) usual or average pain, and 4) pain subscales within PROMs (e.g. KOOS pain). Furthermore, we extracted data on health-related quality of life (HRQoL), kinesiophobia, anxiety and depression, self-efficacy, return to play/work, and self-reported recovery (e.g., GROC). For each eligible follow-up (≥12 months), we extracted endpoint scores, change-from-baseline scores, standard deviations or other measures of dispersion, and sample sizes. If multiple timepoints were reported (e.g. 12 and 24 months), we extracted all available data. If multiple pain/function outcomes were available, data were extracted in accordance with the predefined hierarchy.

### Risk of Bias

Risk of bias was assessed independently by at least two reviewers (KDL, SG, MBB, AA, LBS, NP, CaV, CLS, and SKJ) using the Quality In Prognosis Studies (QUIPS) tool.^16^ Disagreements were resolved through discussion or, when necessary, adjudicated by a senior reviewer (MSR). The QUIPS tool evaluates six domains (study participation, study attrition, prognostic factor measurement, outcome measurement, study confounding, and statistical analysis and reporting) relevant to prognosis research, however, in accordance with our protocol, the domain on prognostic factor measurement was not assessed, as this review focused on long-term outcome prognosis rather than identifying prognostic factors. Each domain was rated as having low, moderate, or high risk of bias based on predefined signalling questions.^16^ To ensure consistent interpretation and minimise subjectivity, a calibration exercise was conducted prior to formal assessment. This involved all reviewers independently rating five sample studies, followed by group discussion to align understanding and refine operational criteria for each domain. Risk of bias assessments were managed and documented in Excel.

### Statistical Analysis

Study heterogeneity was assessed through visual examination of the data extraction table on details related to outcomes and endpoints. In cases were pooling of either outcomes or endpoints was not homogeneous, data was summarised narratively. Meta-analysis was conducted when ≥2 studies reported the same outcome using sufficiently comparable definitions, time points, and statistical formats. For continuous outcomes (e.g., knee pain and self-reported knee function), we extracted change scores where available, or endpoint scores when change scores were not reported. Where necessary, change scores were converted to standardised mean differences (SMD) with 95% confidence intervals (CI) to allow pooling across measurement instruments. We included both randomised and non-randomised longitudinal studies and pooled data regardless of treatment allocation, as the aim was to estimate overall prognosis rather than comparative treatment effects. For randomised trials, outcome data from all study arms were included, with each arm treated as an independent cohort when extracting longitudinal outcome data. Between-study heterogeneity was assessed using the I² statistic, with thresholds of low (0%–25%), moderate (26%–74%), and high (≥75%) heterogeneity.^17^ Where heterogeneity was moderate to high, we used a random-effects model (DerSimonian–Laird estimator), otherwise, a fixed-effect model was applied.^18^ For studies reporting binary outcomes (e.g., treatment success), we calculated pooled proportions and their 95% CI, with success defined as global improvement on GROC or equivalent transition scales. Where appropriate, we conducted meta-regression analyses to explore whether prognosis varied by study-level covariates, such as age, baseline pain severity, or symptom duration. Meta-regressions were conducted using restricted maximum likelihood estimation, and reduction in between-study variance (τ²) was used to evaluate explanatory power.^19^ To assess certainty in the body of evidence for each meta-analysed outcome, we applied the GRADE (Grading of Recommendations, Assessment, Development and Evaluation) approach.^20,21^ Where possible, this included pain and function outcomes across all hierarchy levels and follow-up points. Certainty was downgraded based on five domains: risk of bias (based on QUIPS assessments), inconsistency (heterogeneity across studies), indirectness (differences in population, outcome, or context), imprecision (wide CIs or small sample sizes), and publication bias (funnel plot asymmetry or Egger’s test). Two reviewers (KDL, EM, or GV) independently rated each outcome, with disagreements resolved by consensus. All statistical analyses were performed using R (R Foundation for Statistical Computing, Vienna, Austria), including the ‘meta’²¹ and ‘metafor’ packages, and statistical significance set at p<0.05.

### Equity, diversity, and inclusion statement

This study was conducted by a multidisciplinary research team comprising 12 authors (6 women, 6 men), with representation from physiotherapy, medicine, translational medicine, biology, and information science. The team spans a range of academic stages, including early-career researchers without a PhD, mid-career researchers at postdoctoral or above, and senior academics including two full professors. Authors are affiliated with institutions across three countries including Denmark, Brazil, and Turkey.

## RESULTS

### Search results

The database searches, including all updated searches, identified a total of 29,104 records. After removing 5,975 duplicates, 24,066 titles and abstracts were screened. Of these, 235 full-text articles were assessed for eligibility, and 42 studies met the inclusion criteria.^12,13,22–61^ The full selection process is illustrated in **Figure 1**.

### Characteristics of included studies

At baseline, included studies yielded a cumulative total of 3,230 individuals with PFP. Most participants were female (62.3%), although 2 studies did not report sex. Mean age was 24 years, and the most common age range was 20-30 years (27 studies, 64.3%), followed by <19 years (9, 21.4%), and 31-45 (4, 9.5%). Mean age values were not reported in 2 studies. Across the 42 studies, 23 were randomised controlled trials (54.8%), 13 were interventional studies (31%) and 6 (14.3%) were prospective observational cohort studies. Most common follow-up period was 12 months (+/- 3 months) in 27 studies (64.3%), followed by ≥60 months in 9 (21.4%); 24 (+/- 3 months) in 7 studies (16.7%), and 29- and 36-months follow-ups were investigated in one study each (2, 4.8%). Three studies investigated more than one follow-up period. ^37,50,51^ A summary of data on the characteristics, intervention/control type, and outcomes of the include studies is presented in **Table 1**.

### Outcome measures

Pain intensity was the most frequently assessed outcome across included studies. The VAS was the most commonly used pain measure (n = 20, 47.6%), followed by the NPRS (n = 8, 19%); four studies did not specify the pain measurement tool used. Function and participation-related outcomes were assessed using a range of PROMs, most commonly the AKPS (n = 17, 26.2%), followed by measures of sport participation (n = 8, 18%), KOOS (n = 7, 16.7%), and the Functional Index Questionnaire (FIQ) (n = 4, 9.5%). Additional outcomes included global improvement or recovery (e.g. GROC or post-operative grading), psychosocial measures, clinical performance tests, and health care utilisation. A complete overview of all outcome measures is provided in **Supplementary Appendix 2**.

### Bias and certainty of evidence

Overall, 8 studies were judged as having low risk of bias, 16 studies were judged as moderate risk of bias, and 18 studies were judged high risk of bias. The most common high risk of bias domains were statistical analysis and reporting (n = 10) and study attrition (n = 9). The most common moderate risk of bias domains was study attrition (n = 18) and study confounding (n = 16). The most common low risk of bias domains was outcome measurement (n =13) and study confounding (n = 6). **Table 2** shows the QUIPS assessments on risk of bias from all individual included studies.

**Table 2.**
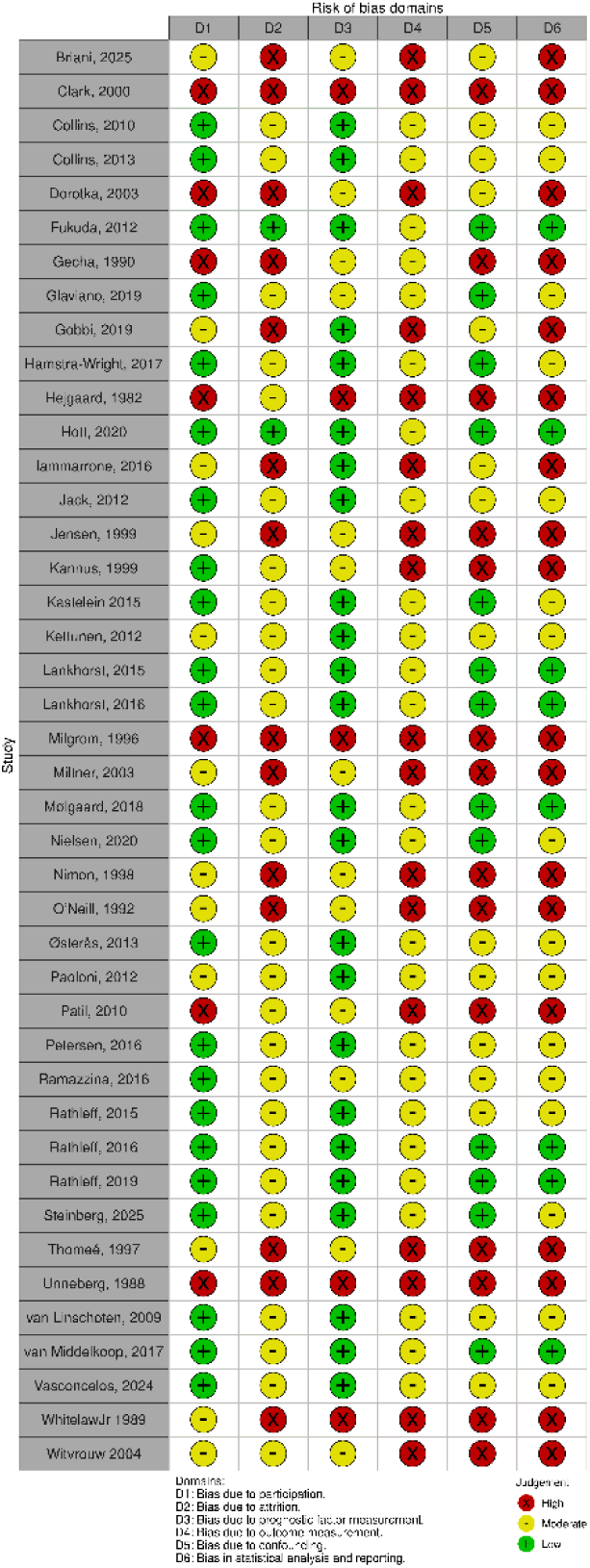
Individual Risk of bias assessment across included studies. Table text: Risk of bias assessments were conducted using the QUIPS framework and visualised using the robvis tool (Risk-Of-Bias VISualization).^62^ To align with the focus of this review on overall prognosis rather than prognostic factors, the original QUIPS domain assessing prognostic factor measurement was removed. Accordingly, domains were re-specified as follows: D1 = study participation, D2 = study attrition, D3 = outcome measurement, D4 = study confounding, D5 = statistical analysis and reporting, and D6 = overall judgement. Colour coding reflects low, moderate, or high risk of bias within each domain.

According to the GRADE approach, the certainty of evidence supporting the estimated effects on pain-related outcomes and self-reported knee function ranged from very low to moderate. For secondary outcomes, the certainty of evidence ranged from low to moderate. These ratings primarily reflect concerns regarding risk of bias, inconsistency, and imprecision across studies. Full GRADE ratings and justifications are available in **Supplementary Appendix 3**.

### Main analysis (Meta-analysis)

#### Primary outcomes

A meta-analysis of seven studies (n = 857) demonstrated a significant reduction in worst pain intensity at the 12-month follow-up (SMD 1.36; 95% CI 0.85 to 1.86; I²=95.09; p<0.001; **Figure 2**), supported by a moderate certainty of evidence. ^12,24,28,32,51,59,63^ Consistent findings were observed for pain during activity, with six studies (n = 406) showing a significant reduction at 12 months (SMD 1.36; 95% CI 0.61 to 2.11; I²=96.14; p<0.001; **Figure 2**), although the certainty of evidence was low. ^23,26,43,57,58^ In contrast, pooled data from eight studies (n = 616) investigating usual pain did not show a significant reduction at this time point (SMD 2.44; 95% CI −0.23 to 5.12; I²=99.81; p=0.07; **Figure 2**), with low certainty of evidence. ^24,28,32,37,39,47,50,59^ In addition, a meta-analysis of five studies (n = 584) investigating resting pain, showed an overall significant reduction from baseline to the 12-month follow-up (SMD 0.91; 95% CI: 0.75 to 1.08; I² = 34.14%; p<0.001; **Figure 2**), supported by moderate certainty of evidence. ^12,39,43,46,58^

**Figure 2.**
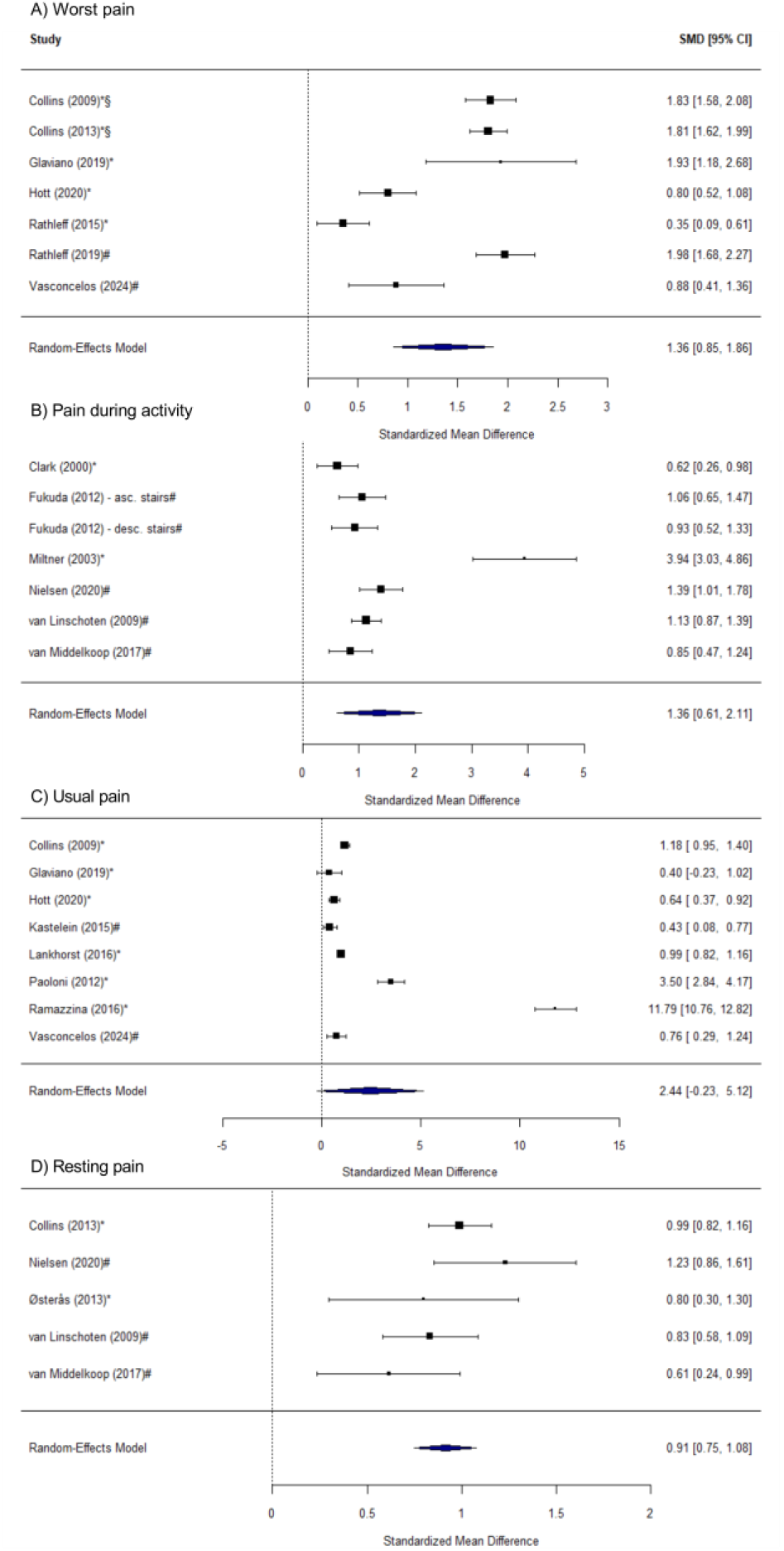
Meta-analysis of A) Worst pain; B) Pain during activity; C) Usual pain; and D) Resting pain from baseline to 12 months follow-up. *: measured with Visual Analogue Scale (VAS); #: measured with Numeric Pain Rating Scale (NPRS).

At the 24-month follow-up, three studies (n = 268) evaluated usual pain, with the pooled analysis indicating no significant improvement (SMD 4.44; 95% CI −3.05 to 11.92; I²=99.86%; p=0.25; **Figure 3**) and the certainty of evidence was very low. ^22,50,51^

**Figure 3.**
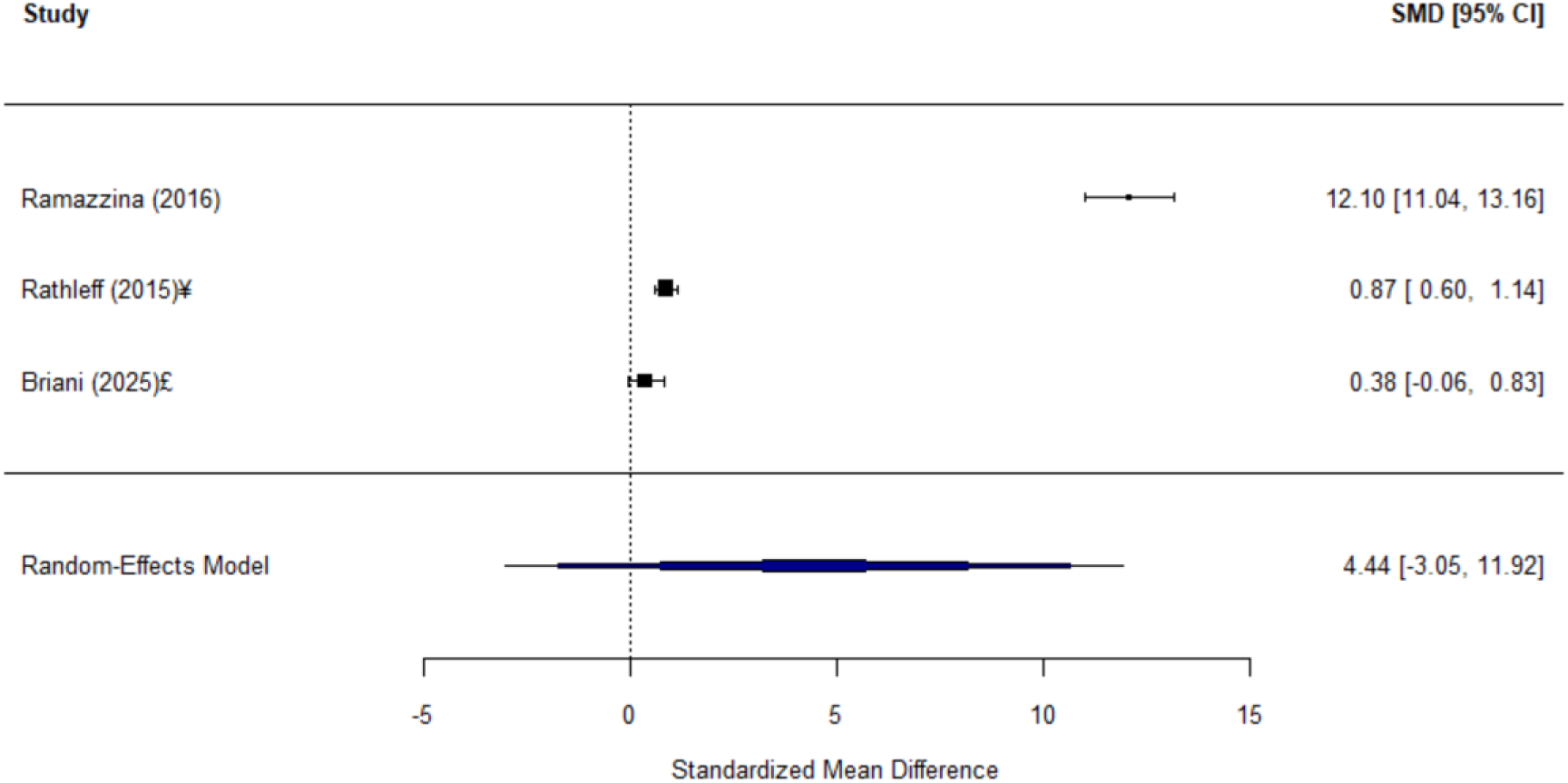
Meta-analysis of pain intensity from baseline to 24 months follow-up. ¥: pain intensity related to last week; £: pain intensity related to previous month.

For extended follow-up (≥60 months), meta-analysis of three studies (n = 157) showed a significant reduction in worst pain intensity (SMD 1.06; 95% CI 0.50 to 1.62; I²=82.17; p<0.001; **Figure 4**), although the evidence was very low. ^34,39,61^ Similarly, pain during activity significantly improved at ≥60 months follow-up based on pooled data from three studies (n = 702) (SMD 0.92; 95% CI 0.76 to 1.07; I²=54.12%; p<0.001; **Figure 4**), with a low certainty of evidence. ^38,39,61^ Usual pain also showed a significant reduction over the same period, as reported in three studies (n = 156) (SMD 1.11; 95% CI 0.88 to 1.34; I²=0%; p<0.001; **Figure 4**), also supported by low-certainty evidence. ^34,39,61^

**Figure 4.**
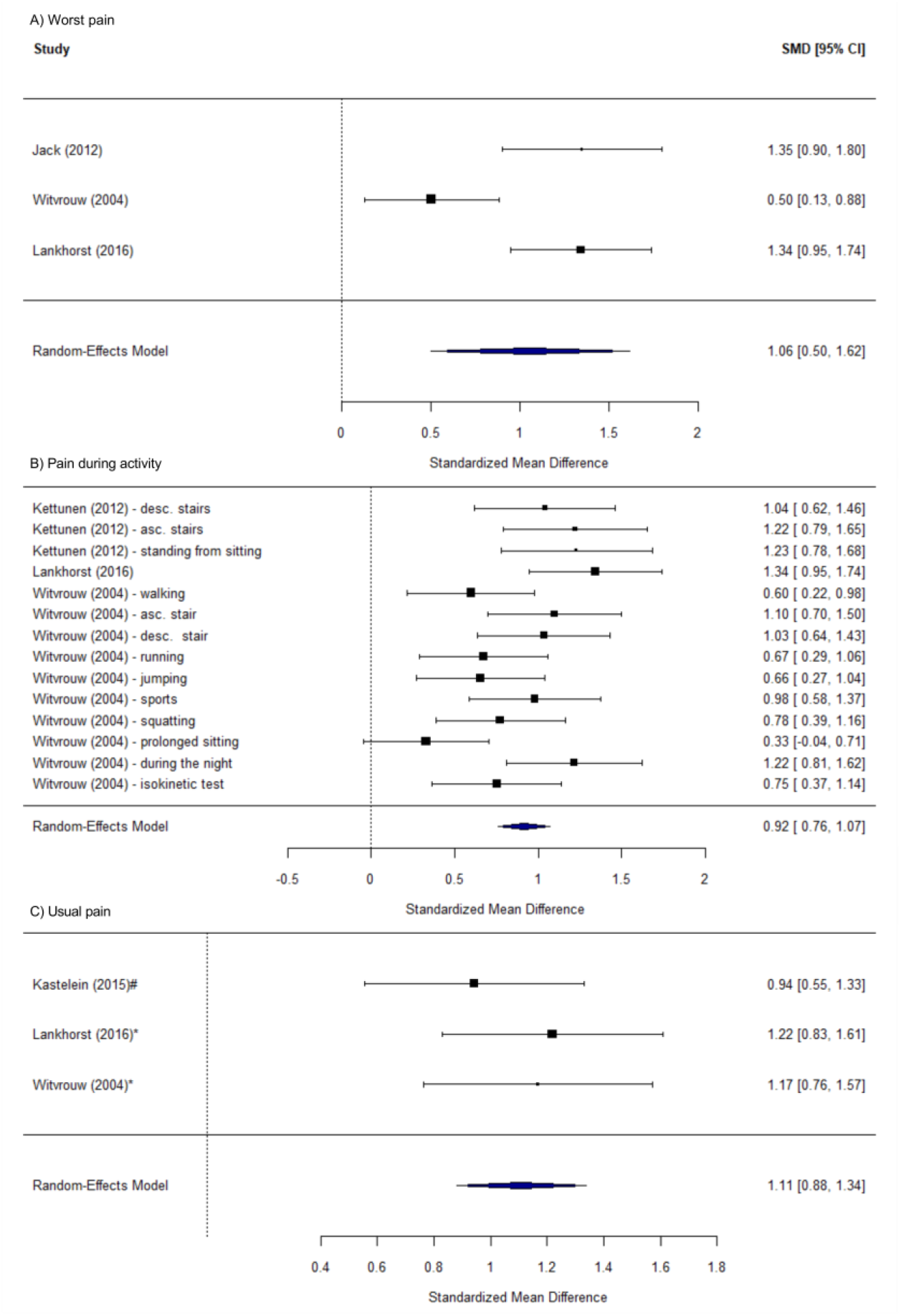
Meta-analysis of A) Worst pain; B) Pain during activity; and C) Usual pain from baseline to ≥60 months follow-up. *: measured with Visual Analogue Scale (VAS); #: measured with Numeric Pain Rating Scale (NPRS).

For function-related self-reported outcomes, at the 12-month follow-up period, meta-analysis of eleven studies (n = 1,015) using the AKPS demonstrated a significant improvement from baseline to the 12-month follow-up (MD: 14.60; 95% CI 11.60 to 17.61; I²=93.41%; p<0.001; **Figure 5**), although the certainty of evidence was low. ^12,24,26,29,32,43,57–59^ Pooled data from three studies (n = 509) using the FIQ showed a significant improvement over the same period (MD 3.33; 95% CI: 2.46 to 4.20; I² = 90.96%; p<0.001; **Figure 5**), also supported by low certainty of evidence. ^12,24,46^ Two studies (n = 114) assessing physical function with the WOMAC also demonstrated a significant improvement from baseline to the 12-month follow-up (MD –7.73; 95% CI: -10.36 to – 5.10; I² = 0%; p<0.001; **Figure 5**), with low certainty of evidence.^23,37^

**Figure 5.**
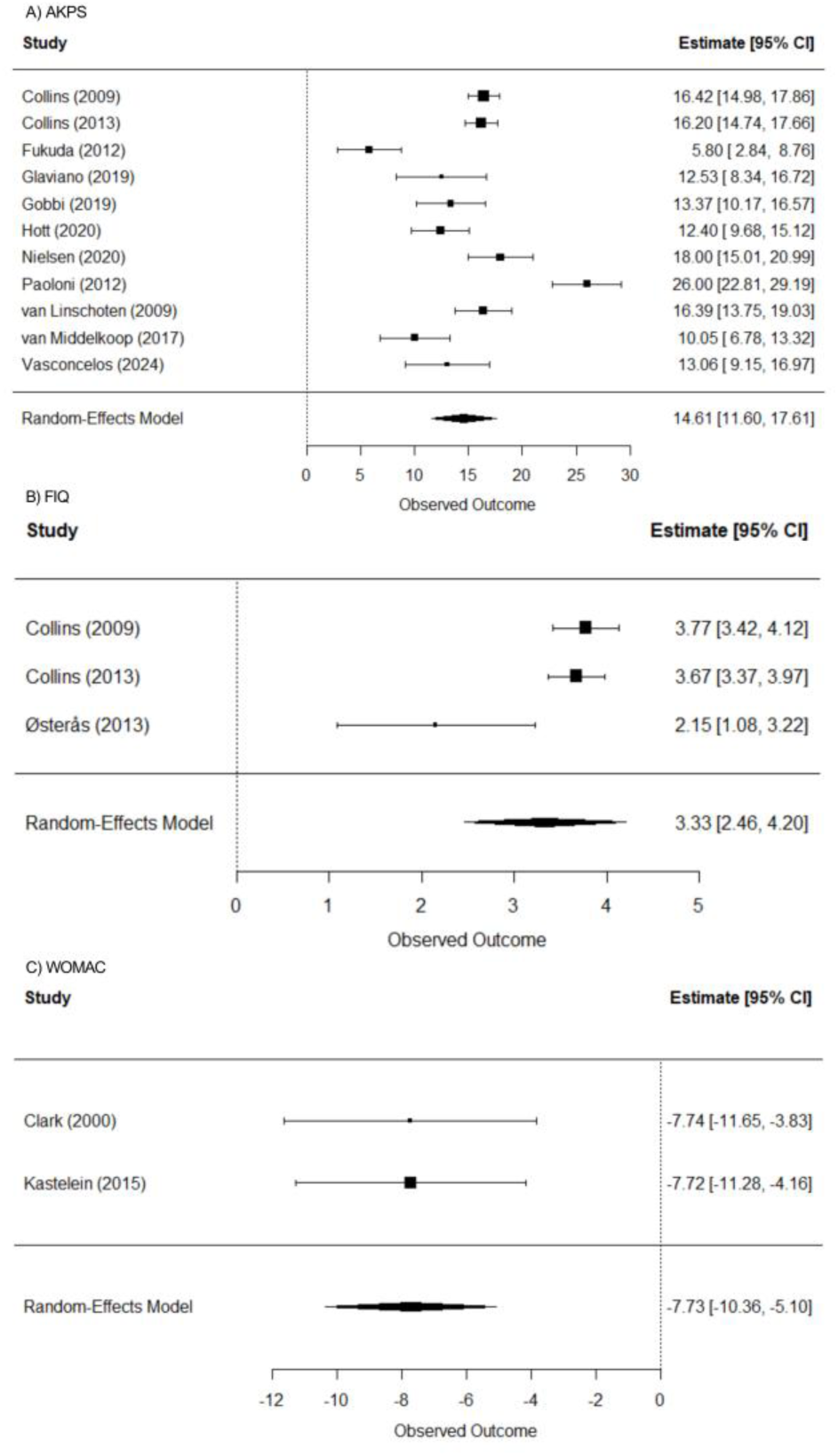
Meta-analysis of physical function measured using A) Anterior Knee Pain Scale (AKPS); B) Functional Index Questionnaire (FIQ); C) Western Ontario and McMaster Universities Osteoarthritis Index (WOMAC) from baseline to 12 months follow-up.

At the extended follow-up (≥60 months), four studies (n = 195) assessed physical function using the AKPS and Lysholm scores, and the meta-analysis showed a significant overall improvement (SMD –1.75; 95% CI: -3.07 to -0.44; I² = 97.07%; p= 0.01; **Figure 6**); however, this finding was based on very low-certainty evidence. ^34,37–39^

**Figure 6.**
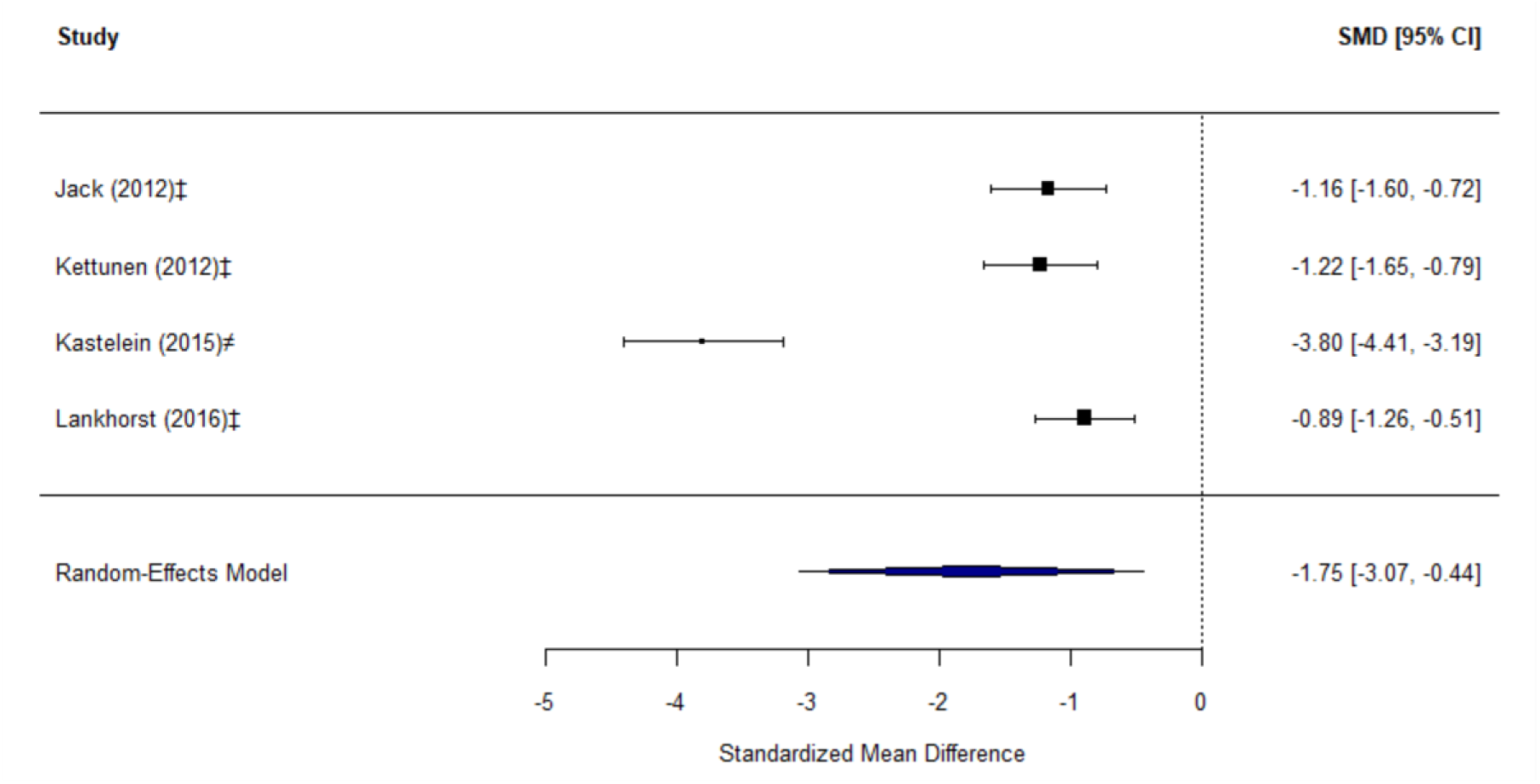
Meta-analysis of physical function from baseline to ≥60 months follow-up. ‡: Anterior Knee Pain Scale (AKPS); ≠: Lysholm Knee Scoring Scale (LKS).

#### Secondary outcomes

Three studies (n = 192) evaluated all five subscales of the KOOS at the 12-month follow-up. ^42,51,58^ Meta analysis of the pooled data showed significant improvements from baseline across all domains (p<0.01), with effect sizes ranging from small to large: Pain (MD = 10.48; 95% CI 6.93 to 14.02; I² = 67.81 %), Symptoms (MD = 4.71; 95% CI 2.12 to 7.30; I² = 57.58 %), Activities of Daily Living (ADL) (MD = 6.78; 95% CI 2.52 to 11.08; I² = 78.14%), Sport (MD = 13.31; 95% CI 3.86 to 22.75; I² = 90.54%), and Quality of Life (QoL) (MD = 7.60; 95% CI 5.94 to 9.25; I² = 0%) (**Supplementary Appendix 4**). The certainty of evidence was moderate for all domains, except for KOOS ADL and KOOS Sport, which were supported by low-certainty evidence.

Additionally, eight studies (n = 1,078) assessed patients’ perception of improvement using the GROC scale and the meta-analysis revealed a significant overall effect of improvement at the 12-month follow-up of 0.54 (95% CI: 0.38 to 0.68; I² = 93.6%; p<0.001; **Supplementary Appendix 4**), supported by a moderate certainty of evidence. ^12,24,37,39,51,57,58^ In contrast, at extended follow-up (≥60 months), pooled data from three studies (n = 160) also showed that the pooled proportion of participants reporting improvement was 0.54 (95% CI: 0.43 to 0.65; I² = 52%; p= 0.12; **Figure 8**), but without a significant effect and with low certainty of evidence. ^37–39^

#### Meta-regression

Among the potential study-level covariates, including age, baseline pain severity, and symptom duration, sufficient data were available to perform meta-regression analyses only for age. At the 12-month follow-up, no significant association was observed between age and improvements in physical function measured by the AKPS (β = −0.03; 95% CI −0.10 to 0.03; p = 0.29; **Figure 7**).

**Figure 7.**
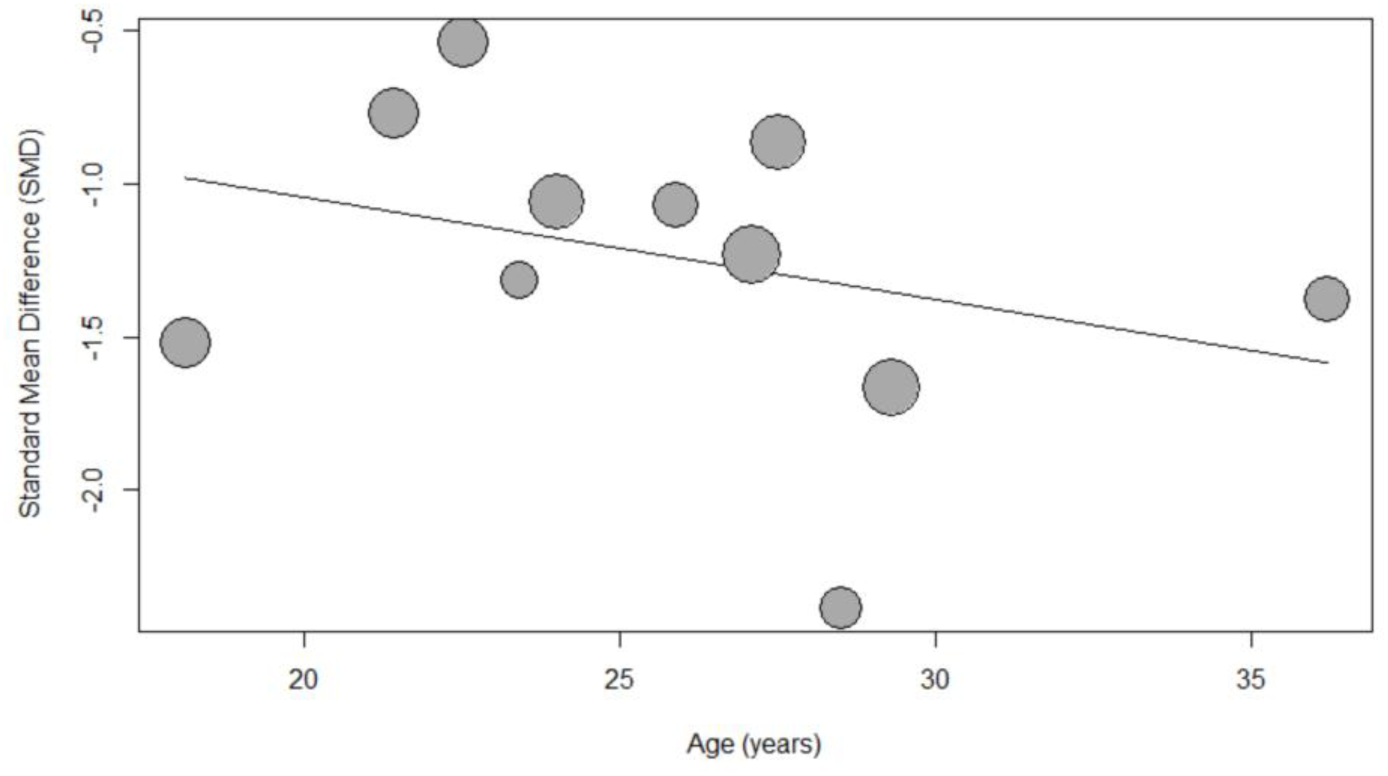
Meta-regression using age as a moderator for physical function from baseline to 12-month follow-up.

## DISCUSSION

### Principal findings

This systematic review and meta-analysis provide the most comprehensive synthesis to date of long-term prognosis (≥12 months) in adolescents and adults with PFP. Overall, pooled analyses suggest that pain and self-reported function improve over time, particularly within the first 12 months. Reductions were observed for worst pain, pain during activity, and resting pain at 12 months, alongside improvements in self-reported function measured by AKPS, FIQ, WOMAC, and KOOS subscales. However, usual pain did not consistently demonstrate statistically significant reduction at 12 months and was characterised by substantial heterogeneity. At extended follow-up (≥60 months), pooled estimates continued to suggest a reduction in pain and in function, but these findings were based on fewer studies, often with very high heterogeneity and lower certainty of evidence. Perceived global improvement showed a moderate effect at 12 months but was less consistent beyond five years. Importantly, a substantial proportion of individuals continued to report persistent symptoms in narrative and proportion-based analyses, indicating that improvement at the group level does not equate to full recovery for all individuals. Meta-regression analyses demonstrated no association between age and functional improvement at 12 months; however, older age was associated with larger improvements in activity-related pain at ≥60 months, although this finding should be interpreted cautiously due to limited data and residual heterogeneity. Certainty of evidence ranged from very low to moderate across outcomes, primarily downgraded due to risk of bias, inconsistency, and imprecision. Collectively, these findings suggest that while the long-term prognosis of PFP is generally favourable at a group level, clinically meaningful symptom persistence remains common, and substantial variability exists across individuals and timeframes.

### Comparison with existing literature

The present findings reinforce and extend a growing body of literature indicating that PFP is often characterised by a condition that presents with a variable course rather than a uniformly self-limiting condition.^12,13,52,53^ Although group-level reduction in pain and improvement in function were observed at 12 months and beyond, narrative data and proportion-based outcomes indicate that a clinically meaningful subgroup continues to experience persistent symptoms.^23,36,44,52,54,56,61^ This supports the view that PFP often follows a prolonged and variable trajectory rather than complete resolution. Previous research has also highlighted that individuals with persistent PFP may experience broader pain and psychosocial features, including multi-site pain, altered pain processing, and psychological factors such as pain catastrophising and fear of movement.^6,7,64^ While these factors were not directly examined in the present review, they may help explain the variability in long-term symptom trajectories observed across studies. Taken together, these findings suggest that the long-term prognosis of PFP may reflect more than local mechanical factors and may instead involve biopsychosocial contributors that influence recovery trajectories. Within this broader prognostic context, previous work by Winters et al. (2021) demonstrated that exercise therapy, education, and orthoses are superior to wait-and-see approaches at 12 months.^8^ However, the present findings indicate that even when evidence-based interventions are provided, residual symptoms remain common in individuals with PFP, reinforcing the need to distinguish between relative treatment efficacy and absolute long-term recovery. In addition, our results extend the evidence and gap map by Neal et al. (2025), which highlighted the substantial heterogeneity in outcome measures, populations, and timepoints across PFP prognosis studies.^10^ By quantitatively synthesising long-term pain and function outcomes, the present review addresses one of the key knowledge gaps identified in that map and provides clearer estimates of trajectories beyond the 12-month timeframe traditionally emphasised in clinical trials.

In this context, consensus statements have emphasised the importance of standardised terminology, outcome reporting, and recognition of potential progression toward patellofemoral osteoarthritis.^2,65^ The substantial heterogeneity observed in the present analyses mirrors the variability in outcome selection and reporting previously identified in the field. This underscores the continued need for harmonised outcome frameworks and long-term follow-up in future PFP research.

### Clinical Implications

These findings have several implications for clinical practice. First, the observation of group-level improvement alongside persistent symptoms in a substantial subgroup reinforces the need for realistic prognostic communication. Contemporary clinical practice guidelines for PFP emphasise exercise therapy, education, and load management as first-line treatments, yet they also acknowledge the variability in individual response.^66,67^ The present synthesis suggests that clinicians should view recovery not as a binary outcome but as a trajectory that unfolds over time and varies from person to person, while acknowledging that a meaningful proportion of individuals will continue to experience residual or recurrent symptoms. Second, in light of emerging evidence highlighting the contribution of psychological factors and altered nociceptive processing, together with the differential symptom trajectories observed in the present study, these findings underscore the need for multidimensional outcome monitoring and a biopsychosocial informed management approach. Consistent with broader musculoskeletal guidance advocating multimodal, person-centred care that addresses behavioural and psychosocial contributors (NICE Guideline NG193, 2021), clinicians managing PFP should assess and monitor multiple symptom domains rather than relying on single pain metrics, and tailor interventions to reflect the multidimensional nature of persistent pain. Third, the persistence of symptoms in a subset of individuals highlights the importance of long-term self-management strategies and load adaptation rather than short-term symptom elimination. Applying these principles to PFP management may support sustained participation in sport while reducing symptom exacerbation.^68^ Finally, the limited prognostic value of available study-level factors, alongside the lack of clear associations in meta-regression, suggests that clinicians should be cautious in making firm predictions about individual long-term outcomes. Instead, clinical encounters should prioritise shared decision-making, collaborative goal setting, and iterative review of progress, enabling treatment plans to adapt to the patient’s changing needs, preferences, and contextual factors.^69–71^

In this context and introduced earlier, these findings also help address questions that patients frequently ask in clinical settings: “*When will my pain go away*?” and “*When will I be able to return to sport*?”. ^14^ The present synthesis suggests that, on average, improvements in pain and self-reported function occur within the first 12 months, indicating that many individuals experience meaningful symptom reduction during this period. However, the persistence of symptoms in a substantial proportion of individuals highlights that recovery timelines are highly variable and that complete symptom resolution cannot be guaranteed within a fixed timeframe. Consequently, clinicians should communicate that return to previous activities or sport is often possible through progressive load management and symptom-guided activity modification rather than waiting for complete pain elimination. Framing recovery as a gradual and individualised process may help align patient expectations with the typical trajectory observed in longitudinal studies.

### Limitations

Several limitations should be considered when interpreting these findings. First, substantial between-study heterogeneity was observed across most pooled analyses, with I² values frequently indicating high inconsistency. Although random-effects models were applied where appropriate, such heterogeneity reduces the precision of pooled estimates and suggests that the summary effects should be interpreted as average trajectories across diverse clinical contexts rather than precise predictions for individual patients. The heterogeneity likely reflects variation in participant characteristics, symptom duration, interventions received, outcome measures, and follow-up intervals, as well as differences in study quality.

Second, risk of bias was moderate to high in a considerable proportion of included studies, particularly in domains related to attrition and statistical analysis and reporting. These methodological limitations contributed to downgrading the certainty of evidence and may have influenced effect estimates. While sensitivity analyses were not consistently feasible across all outcomes due to limited numbers of studies per timepoint, the potential impact of bias cannot be excluded.

Third, outcome reporting was highly variable. Multiple pain constructs were measured using different instruments and definitions, necessitating conversion to standardised mean differences. Although this approach enabled quantitative synthesis across heterogeneous measures, it limits direct clinical interpretability and may obscure nuances between specific pain domains. In addition, incomplete reporting of covariates restricted the scope of meta-regression analyses; only age could be examined with sufficient data. Study-level meta-regression is inherently limited in its ability to account for individual-level confounding and should be interpreted as exploratory rather than causal.

Finally, the inclusion of both randomised and non-randomised longitudinal studies, pooled irrespective of treatment allocation, aligns with the objective of estimating overall prognosis but introduces additional clinical variability. Moreover, several studies did not report sufficient data for inclusion in quantitative synthesis at specific timepoints, and extended estimates (≥60 months) were based on relatively few studies, reducing confidence in those findings.

Collectively, these limitations indicate that the pooled estimates should be interpreted as indicative of general trends rather than precise prognostic predictions. While the direction of effect was broadly consistent across outcomes and timepoints, the substantial heterogeneity, risk of bias in several studies, and limited data at extended follow-up periods reduce certainty in the magnitude of effect. Consequently, the findings are best viewed as an aggregate representation of long-term trajectories in PFP rather than definitive estimates applicable to all clinical settings or patient subgroups.

### Implications for future research

Future research should prioritise high-quality prospective studies with harmonised outcome measures and predefined long-term follow-up intervals to improve comparability across cohorts. Adoption of core outcome sets, and standardised reporting frameworks would enhance the interpretability of pooled analyses and reduce heterogeneity. There is also a need for trajectory-based and individual participant data analyses to identify clinically meaningful subgroups and clarify the determinants of persistent versus resolving symptoms. Finally, future studies should integrate multidimensional assessments, including psychosocial and pain-processing measures, to better characterise mechanisms underlying long-term prognosis and to inform stratified or precision-based care models in patellofemoral pain.

## Conclusion

Given the high prevalence and long-term burden of patellofemoral pain in adolescents and adults, clinicians require robust evidence to inform prognostic communication and management strategies. This systematic review and meta-analysis demonstrate that, at a group level, pain and self-reported function generally improve over time, particularly within the first 12 months. However, substantial heterogeneity and persistent symptoms in a meaningful subgroup indicate that recovery is not universal and that long-term trajectories are highly variable. Meta-regression analyses were limited and identified no consistent moderators beyond an exploratory association with age at 12-month follow-up. Collectively, these findings suggest that while the overall prognosis of PFP is moderately favourable, persistent and variable recurrent symptoms remain common. This highlights the need for realistic patient communication, multidimensional management approaches, and further research into stratified long-term care models, including strategies to differentiate patients who recover from those who experience prolonged symptom, as well as targeted education to support patients in learning to manage their pain.

## Data Availability

All data produced in the present study are available upon reasonable request to the authors

## Acknowledgement

The authors would like to thank Dr. Marinus Winters for support during the development of the literature search. Furthermore, the authors would like to thank Assistant Professor, PhD, Sinead Holden for the contributions in the conceptualization and methodology stage.

## Patient and public involvement

Patients and/or the public were not involved in the design, or conduct, or reporting, or dissemination plans of this research.

