## Supplementary material for "Long-term prognosis of patellofemoral pain in adolescents and adults: A systematic review with meta-analysis and meta-regression": Table 1

Table 1. Characteristics of included studies.

| **Author,**  **year** | **Country** | **Study design** | **Setting**  **Recruitment** | **Follow-up** | **N** | **Participants characteristics** | **Type of intervention** | **Outcomes** |
| --- | --- | --- | --- | --- | --- | --- | --- | --- |
| Briani,  2025 | Brazil | Prospective cohort | Secondary | 25 months | 39 | Mean age, years: 22.9  Female/Male, n: 31/8  Mean BMI, kg/m^2^: 24.1  Mean symptom duration, mths: 49.3 | NA | Usual pain (VAS) |
| Clark,  2000 | United Kingdom | RCT | Primary and secondary | 12 months | BA: 81  FU: 49 | Mean age, years: 30  Female/Male, n: 36/45  Mean BMI, kg/m^2^: 25  Mean symptom duration, mths: NR | 1) Exercise + tape  2) Exercise  3) Tape  4) Education | Pain during activity (VAS)  Patient-reported knee function (WOMAC)  Patient satisfaction  Depression (HAD)  Persistent symptoms |
| Collins,  2009 | Australia | RCT | Community setting | 12 months | BA: 179  FU: 171 | Mean age, years: 29.3  Female/Male, n: 100/79  Mean BMI, kg/m^2^: 24.8  Mean symptom duration, mths: 41.3 | 1) Foot orthoses  2) Flat inserts  3) Physiotherapy (PT)  4) Foot orthoses + PT | Worst pain activity-related (VAS)  Patient-reported knee function (AKPS; FIQ)  Improvement perception (GROC) |
| Collins,  2013 | Australia | RCT | Primary care and community setting | 12 months | 310 | Mean age, years: 27.1  Female/Male, n: 184/126  Mean BMI, kg/m^2^: 24  Mean symptom duration, mths (n): 1-2 mths = 58; 2-6 mths = 56; 6-12 mths = 49; > 12 = 143 | 1) Australian RCT: foot orthoses; flat shoe inserts; PT; foot orthoses + PT  2) Dutch trial: exercise therapy; usual care | Worst pain activity-related (VAS)  Resting pain (VAS)  Patient-reported knee function (AKPS; FIQ)  Improvement perception (GROC) |
| Dorotka,  2003 | Austria | Interventional study | Military Hospital | 24 months | 58 | Mean age, years: 21.6  Female/Male, n: 0/58  Mean BMI, kg/m^2^: 23.6  Mean symptom duration, mths: 31.8 | Exercise + 400mg of sodium chondroitin sulphate three times daily. | Pain intensity  Persistent symptoms |
| Fukuda,  2012 | Brazil | RCT | Secondary | 12 months | BL: 54  FU: 49 | Mean age, years: 22.5  Female/Male, n: 0/54  Mean BMI, kg/m^2^: 24.1  Mean symptom duration, mths: 22.1 | 1) Knee exercise  2) Knee + hip exercise | Pain ascending and descending stairs (NPRS)  Patient-reported knee function (AKPS; LEFS)  Functional tests |
| Gecha,  1990 | United States | Interventional study | NR | 29 months | BL: 45  FU: 38 | Mean age, years: 17  Female/Male, n: NR  Mean BMI, kg/m^2^: NR  Mean symptom duration, mths: NR | Surgery: lateral retinacular release | Patient satisfaction  Postoperative grading |
| Glaviano,  2019 | United States | RCT | Secondary | 12 months | BL: 21  FU: 19 | Mean age, years: 23.4  Female/Male, n: 16/5  Mean BMI, kg/m^2^:  Mean symptom duration, mths: 24.7 | 1) Patterned Electrical Neuromuscular Stimulation + PT rehab  2) Sham + PT rehab | Worst pain (VAS)  Patient-reported knee function (AKPS; ADLS) |
| Gobbi,  2019 | Brazil | RCT | Hospital | 12 months | 25 | Mean age, years: 36.2  Female/Male, n: NR  Mean BMI, kg/m^2^: NR  Mean symptom duration, mths: NR | 1) Pulsed Signal Therapy (PST)  2) Control: Sham PST | Patient-reported knee function (AKPS) |
| Hamstra-Wright,  2017 | United States | RCT | University | 12 and 24 months | BL: 157  FU: 78 | Mean age, years: 28.4  Female/Male, n: 105/52  Mean BMI, kg/m^2^: NR  Mean symptom duration, mths: 25.5 | 1) Hip exercise  2) Knee exercise | Injury recurrence |
| Hejgaard,  1982 | Denmark | RCT | Hospital and clinic | 12 months | 42 | Mean age, years: 30  Female/Male, n: 23/19  Mean BMI, kg/m^2^: NR  Mean symptom duration, mths: NR | 1) Shaving  2) Shaving + displacement | Postoperative grading |
| Hott,  2020 | Norway | RCT | Hospital and clinic | 12 months | BL: 112  FU: 98 | Mean age, years: 27.5  Female/Male, n: 73/39  Mean BMI, kg/m^2^: 26.4  Mean symptom duration, mths: 3-6 mths = 8; 6 - 12 mths = 23; 12 -24 mths = 24; > 14 mths = 57 | 1) Knee exercise  2) Hip exercise  3) Patient education | Worst pain (VAS)  Usual pain (VAS)  Patient-reported knee function (AKPS)  Functional tests  Kinesiophobia (TSK)  Self-Efficacy (KSES)  Quality of life (EQ-5D-5L)  Hip and/or knee muscle strength |
| Iammarrone,  2016 | Italy | RCT | University and clinic | 12 months | 30 | Mean age, years: 22.5  Female/Male, n: 22/8  Mean BMI, kg/m^2^: NR  Mean symptom duration, mths: NR | 1) Pulsed Electromagnetic Fields + home exercise program (HEP)  2) HEP | Pain intensity (VAS)  Patient-reported knee function (VISA; Feller’s Patella Score)  Consumption of medication |
| Jack,  2012 | United Kingdom | Interventional study | Hospital and clinic | 72.6 months | BL: 47  FU: 46 | Mean age, years: 34.4  Female/Male, n: 39/7  Mean BMI, kg/m^2^: NR  Mean symptom duration, mths: NR | Modified tibial tubercle osteotomy + PT rehabilitation | Worst pain (VAS)  Patient-reported knee function (AKPS)  Patient satisfaction |
| Jensen,  1999 | Norway | RCT | Primary care and clinic | 12 months | BL: 75  FU: 70 | Mean age, years: 31  Female/Male, n: 41/29  Mean BMI, kg/m^2^: 23.2  Mean symptom duration, mths: 79 | 1) Acupuncture  2) Control: no treatment | Patient-reported knee function (CRS)  Persistent symptoms  Reported physiotherapy/GP consults |
| Kannus,  1999 | Finland | RCT | Hospital and clinic | ~79.2 months | BL: 49  FU: 45 | Mean age, years: 27  Female/Male, n: 28/25  Mean BMI, kg/m^2^: NR  Mean symptom duration, mths: 16 | 1) Non-operative: exercises + oral NSAIDs  2) Non-operative: exercises + intra-articular injections of placebo (saline solution + lidocaine)  3) Non-operative: exercises + intra-articular injections of glycosaminoglycan polysulfate solution | Pain intensity (VAS)  Patient satisfaction  Patient-reported knee function (LKS; TAS)  Functional tests  Hip and/or knee muscle strength  Clinical tests  Structural imaging findings |
| Kastelein,  2015 | The Netherlands | Prospective Cohort | Primary care and care clinic | 12 and 72 months | BL: 74  FU1: 65  FU2: 45 | Mean age, years: 23.7  Female/Male, n: NR  Mean BMI, kg/m^2^: NR  Mean symptom duration, mths: NR | NA | Usual pain (NPRS)  Patient-reported knee function (WOMAC; LKS)  Improvement perception (GROC) |
| Kettunen,  2012 | Finland | RCT | Hospital and clinic | 60 months | BL: 56  FU: 44 | Mean age, years: 28.4  Female/Male, n: 35/21  Mean BMI, kg/m^2^: 24  Mean symptom duration, mths: 50 | 1)Arthroscopy + home exercise  2) Control: home exercise alone | Pain ascending and descending stairs (VAS)  Pain standing from sitting (VAS)  Patient-reported knee function (AKPS)  Improvement perception (GROC) |
| Lankhorst,  2015 | The Netherlands | RCT | Primary care and sports medicine settings | 12 months | BL: 131  FU: 117 | Mean age, years: 24  Female/Male, n: 84/47  Mean BMI, kg/m^2^: 23.1  Mean symptom duration, mths (n):2-6 mths = 89; 6-24 mths = 42 | 1) Exercise  2) Control: usual care | Pain intensity (NPRS)  Patient-reported knee function (AKPS)  Improvement perception (GROC) |
| Lankhorst,  2016 | The Netherlands | RCT | Primary care and community setting | 12 months and 5–8 years | BL: 60  FU: 60 | Mean age, years: 26.2  Female/Male, n: 45/15  Mean BMI, kg/m^2^: 23  Mean symptom duration, mths (n): 1-2 mths = 18; 2-6 mths= 11; 6-12 mths = 9; > 12 = 22 | 1) Australian RCT: foot orthoses; flat shoe inserts; PT and foot orthoses + PT  2) Dutch RCT: exercise therapy; usual care | Worst pain (VAS)  Pain during activity (VAS)  Usual pain (VAS)  Patient-reported knee function (AKPS; FIQ; KOOS)  Improvement perception (GROC)  Structural imaging findings |
| Milgrom,  1996 | Israel | Prospective Cohort | Military | 72 months | BL: 60  FU: 50 | Mean age, years: NR  Age, range: 17 to 25  Female/Male, n: 0/60  Mean BMI, kg/m^2^: NR  Mean symptom duration, mths: NR | NA | Pain intensity  Persistent symptoms |
| Miltner,  2003 | Germany | Interventional study | Secondary | 36 months | 27 | Mean age, years: 15.8  Female/Male, n: 24/3  Mean BMI, kg/m^2^: NR  Mean symptom duration, mths: NR | Surgical procedure: patellar decompression | Pain during activity (VAS)  Clinical tests  Patellar pressure |
| Mølgaard,  2018 | Denmark | RCT | Hospital and clinic | 12 months | BL: 40  FU: 32 | Mean age, years: 31.2  Female/Male, n: 28/12  Mean BMI, kg/m^2^: NR  Mean symptom duration, mths: NR | 1) Knee and foot exercise + orthoses  2)Knee targeted exercises (standard practice) | Patient-reported knee function (KOOS)  Patient satisfaction  Consumption of medication |
| Nielsen,  2020 | Denmark | Interventional study | Community and clinic | 12 months | 65 | Mean age, years: 18.1  Female/Male, n: 52/13  Mean BMI, kg/m^2^: NR  Mean symptom duration, mths: 26 | Patient education + exercise program + shoes/orthoses recommendation | Pain during activity (NPRS)  Resting pain (NPRS)  Patient-reported knee function (AKPS)  Patient satisfaction |
| Nimon,  1998 | United Kingdom | Interventional study | Hospital and clinic | Mean 3.8 years (range: 2-8) and 16.0 years (range: 14-20) | BL: 63  FU1: 54  FU2: 49 | Mean age, years: 15.5  Female/Male, n: 0/63  Mean BMI, kg/m^2^: NR  Mean symptom duration, mths: NR | Non-operatively (not further described) | Persistent symptoms  Participation in sports/activities  Consumption of medication |
| O’Neill,  1992 | United States | Interventional study | Hospital | 12 to 16 months | BL: 34  FU: 30 | Mean age, years: NR  Age range: 10 – 50 years  Female/Male, n: 21/9  Mean BMI, kg/m^2^: NR  Mean symptom duration, mths: NR | Isometric quadriceps strengthening exercises | Pain intensity  Patient satisfaction  Participation in sports/activities  Structural imaging findings  Consumption of medication |
| Østerås,  2013 | Norway | RCT | Primary care and clinic | 12 months | BL: 40  FU: 28 | Mean age, years: 29.9  Female/Male, n: 22/6  Mean BMI, kg/m^2^: NR  Mean symptom duration, mths: 39 | 1) Intervention: high-dose, high-repetition MET exercise therapy  2) Control: ow-dose, low-repetition exercise therapy | Resting pain (VAS)  Patient-reported knee function (FIQ)  Functional tests |
| Paoloni,  2012 | Italy | Interventional study | Hospital | 12 months | 44 | Mean age, years: 28.5  Female/Male, n: 29/15  Mean BMI, kg/m^2^: NR  Mean symptom duration, mths: 13.7 | Patellar taping + specific exercise program | Usual pain (VAS)  Patient-reported knee function (AKPS)  Hip and/or knee muscle strength |
| Patil,  2010 | United Kingdom | Interventional study | School and colleges | Mean 20 months (12-48 months) | 34 | Mean age, years: 17  Female/Male, n: 20/14  Mean BMI, kg/m^2^: NR  Mean symptom duration, mths: NR | Physiotherapy (not specified) | Quality of life (SF-36) |
| Petersen,  2016 | Germany | RCT | Hospital and clinic | 12 months | BL: 156  FU: 130 | Mean age, years: 28  Female/Male, n: 94/36  Mean BMI, kg/m^2^: 23  Mean symptom duration, mths: NR | 1) Intervention:  brace + exercise    2) Control: exercise alone | Pain intensity (VAS)  Patient-reported knee function (AKPS; KOOS)  Improvement perception (GROC)  Consumption of medication |
| Ramazzina,  2016 | Italy | Interventional study | Clinic | 12 and 24 months | BL: 134  FU1: 134  FU2: 130 | Mean age, years: 21.4  Female/Male, n: 85/49  Mean BMI, kg/m^2^: NR  Mean symptom duration, mths: NR | Rehabilitation program (selective muscle strengthening) | Usual pain (VAS)  Patient-reported knee function (CRS)  Hip and/or knee muscle strength |
| Rathleff,  2015 | Denmark | RCT | School | 12 and 24 months | BL: 121  FU1: 110  FU2: 99 | Mean age, years: 17.2  Female/Male, n: 97/24  Mean BMI, kg/m^2^: 21.7  Mean symptom duration, mths: 39 | 1) Exercise therapy + patient education    2) Patient education alone | Worst pain (VAS)  Usual pain (VAS)  Patient-reported knee function (KOOS)  Quality of life (EQ-5D-5L)  Improvement perception (GROC)  Patient satisfaction  Physical activity level (PAS)  Participation in sports/activities  Consumption of medication |
| Rathleff,  2016 | Denmark | Prospective Cohort | School | 24 months | BL: 153  FU: 127 | Mean age, years: 17  Female/Male, n: 126/27  Mean BMI, kg/m^2^: 21.7  Mean symptom duration, mths: 37.5 | NA | Pain intensity (VAS)  Patient-reported knee function (KOOS)  Quality of life (EQ-5D-5L)  Persistent symptoms  Physical activity level (PAS)  Participation in sports/activities |
| Rathleff,  2019 | Denmark | Interventional study | School and community | 12 months | BL: 151  FU: 120 | Mean age, years: 12.6  Female/Male, n: 115/36  Mean BMI, kg/m^2^: 19  Mean symptom duration, mths: 18 | Activity modification and load management | Worst pain (NPRS)  Patient-reported knee function (KOOS)  Quality of life (EQ-5D-Y-5L)  Patient satisfaction  Improvement perception (GROC)  Persistent symptoms  Participation in sports/activities  Consumption of medication |
| Steinberg,  2025 | Israel | Prospective Cohort | Dance school | 12 months | 18 | Mean age, years: 11.9  Female/Male, n: 18/0  Mean BMI, kg/m^2^: 18.7  Mean symptom duration, mths: NR | NA | Persistent symptoms |
| Thomeé,  1997 | Sweden | RCT | Primary | 12 months | BL: 40  FU: 40 | Mean age, years: 20.2  Female/Male, n: 40/0  Mean BMI, kg/m^2^: NR  Mean symptom duration, mths: 43 | 1) Isometric exercises  2) Eccentric exercises | Pain intensity (VAS)  Patient-reported knee function  Patient satisfaction  Functional tests  Participation in sports/activities  Muscle activity  Hip and/or knee muscle strength |
| Unneberg,  1988 | Norway | Interventional study | Hospital and clinic | 3-5 years | 28 | Mean age, years: 23  Female/Male, n: 11/17  Mean BMI, kg/m^2^: NR  Mean symptom duration, mths: NR | Surgical: lateral retinaculum release | Persistent symptoms  Participation in sports/activities  Atrophy |
| van Linschoten, 2009 | The Netherlands | RCT | General practice and clinic | 12 months | BL: 131  BU: 117 | Mean age, years: 24  Female/Male, n: 84/47  Mean BMI, kg/m^2^: 23.1  Mean symptom duration, mths: NR | 1)Intervention: exercise therapy    2) Control: usual care (“wait and see” approach) | Pain during activity (NPRS)  Resting pain (NPRS)  Patient-reported knee function (AKPS)  Improvement perception (GROC) |
| van Middelkoop, 2017 | The Netherlands | Prospective Cohort | Primary care and clinic | 12 months | BL: 64  FU: 50 | Mean age, years: 21.4  Female/Male, n: 35/29  Mean BMI, kg/m^2^: 22.8  Mean symptom duration, mths: 12.6 | NA | Pain during activity (NPRS)  Resting pain (NPRS)  Patient-reported knee function (AKPS; KOOS)  Improvement perception (GROC)  Reported physiotherapy/GP consults |
| Vasconcelos, 2024 | Brazil | RCT | Community and clinic | 12 months | BL: 37  FU: 29 | Mean age, years: 25.9  Female/Male, n: 27/10  Mean BMI, kg/m^2^: 24.9  Mean symptom duration, mths: 58 | 1) Muscle strength and power training program  2) Muscle strength training alone | Worst pain (NPRS)  Usual pain (NPRS)  Patient-reported knee function (AKPS) |
| Whitelaw Jr., 1989 | United States | Interventional study | Community and clinic | 16 months | BL: 85 | Mean age, years: 30  Female/Male, n: 51/34  Mean BMI, kg/m^2^: NR  Mean symptom duration, mths: NR | PT program + NSAIDs medication | Patient-reported knee function  Patient satisfaction |
| Witvrouw,  2004 | Belgium | RCT | University | 60 months | BL: 60  FU: 51 | Mean age, years: 20.3  Female/Male, n: 40/20  Mean BMI, kg/m^2^: NR  Mean symptom duration, mths: 15.2 | 1) OKC exercise program  2) CKC exercise program | Worst pain (VAS)  Pain walking (VAS)  Pain ascending and descending stairs (VAS)  Pain running (VAS)  Pain jumping (VAS)  Pain sports (VAS)  Pain squat (VAS)  Pain prolonged sitting (VAS)  Pain during the nigth (VAS)  Pain isokinetic test (VAS)  Usual pain (VAS)  Patient-reported knee function (AKPS)  Functional tests  Hip and/or knee muscle strength  Persistent symptoms  Participation in sports/activities |

RCT: randomized clinical trial; BA: baseline; FU: follow-up; NA: not applicable; NR: not reported; VAS: Visual Analogue Scale; NPRS: Numeric Pain Rating Scale; WOMAC: Western Ontario and McMaster Universities Osteoarthritis Index; HAD: Hospital Anxiety and Depression Scale; AKPS: Anterior Knee Pain Scale; FIQ: Functional Index Questionnaire; LKS: Lysholm Knee Scoring Scale; KOOS: Knee injury and Osteoarthritis Outcome Score; TAS: Tegner Activity Scale; GROC: Global Rating of Change; LEFS: Lower Extremity Functional Scale; ADLS: Activities of Daily Living Scale; TSK: Tampa Scale for Kinesiophobia; KSES: Knee Self-Efficacy Scale; EQ-5D-5L: EuroQol five-dimension five-level questionnaire; SF-36: 36-Item Short Form Health Survey; EQ-5D-Y-5L: EuroQol five-dimension five-level youth version; PAS: Physical Activity Scale; VISA: Victorian Institute of Sport Assessment; CRS: Cincinnati Rating System; mths: months; OKC: open kinetic chain; CKC: closed kinetic chain.
