## Supplementary Appendices for "Long-term prognosis of patellofemoral pain in adolescents and adults: A systematic review with meta-analysis and meta-regression"

**Supplementary Appendix 1. Search Strategy**

**EMBASE:**

1) "patellofemoral pain syndrome"/exp
2) "anterior knee pain"/exp
3) patell*:ab,ti  OR femoropatell*:ab,ti OR femoro-patell*:ab,ti OR retropatell*:ab,ti OR retro-patell*:ab,ti OR "anterior knee*":ab,ti OR peripatell*:ab,ti OR peri-patell*:ab,ti OR kneecap*:ab,ti
4) pain*:ab,ti OR sore*:ab,ti OR discomfort*:ab,ti OR arthralgia*:ab,ti OR dysfunction*:ab,ti OR injur*:ab,ti OR syndr*:ab,ti OR chondromalac*:ab,ti OR chondropath*:ab,ti
5) "clinical study"/exp OR "controlled study"/exp OR "cohort analysis"/exp OR "observational study"/exp
6) "randomised controlled trial*":ab,ti OR "randomised clinical trial*":ab,ti OR "randomized controlled trial*":ab,ti OR "randomized clinical trial*":ab,ti OR "controlled trial*":ab,ti OR "controlled study*":ab,ti OR "clinical trial*":ab,ti  OR "quasi-randomised":ab,ti OR "quasi-randomized":ab,ti OR "semi-randomised":ab,ti OR "semi-randomized":ab,ti OR "observational stud*":ab,ti OR cohort*:ab,ti OR longitudinal*:ab,ti OR "prospective":ab,ti OR "long term follow-up":ab,ti OR "long-term follow-up":ab,ti OR "prognostic":ab,ti
7) # 1 OR #2
8) #3 AND #4
9) #5 OR #6
10) #7 OR #8
11) #9 AND #10

**MEDLINE via Pubmed**

1) "Patellofemoral Pain Syndrome"[Mesh]
2) "Chondromalacia Patellae"[Mesh]
3) patell*[tiab] OR femoropatell*[tiab] OR retropatell*[tiab] OR "anterior knee*"[tiab] OR peripatell*[tiab] OR "kneecap"[tiab] OR patellofemoral[tiab] OR patello-femoral[tiab]
4) pain*[tiab] OR sore*[tiab] OR discomfort*[tiab] OR arthralgia*[tiab] OR dysfunction*[tiab] OR injur*[tiab] OR syndr*[tiab] OR chondromalac*[tiab] OR chondropath*[tiab]
5) "clinical study"[Publication Type] OR "observational study"[Publication type]
6) "randomised controlled trial*"[tiab]  OR "randomised clinical trial*"[tiab]  OR "randomized controlled trial*"[tiab]  OR "randomized clinical trial*"[tiab]  OR "controlled trial*"[tiab]  OR "controlled study*"[tiab]  OR "clinical trial*"[tiab] OR "quasi-randomised"[tiab]  OR "quasi-randomized"[tiab]  OR "semi-randomised"[tiab] OR "semi-randomized"[tiab]  OR "observational stud*"[tiab] OR cohort*[tiab] OR longitudinal*[tiab] OR "prospective"[tiab]  OR "long term follow-up"[tiab]  OR "long-term follow-up"[tiab] OR "prognostic"[tiab]
7) #1 OR #2
8) #3 AND #4
9) #5 OR #6
10) #7 OR #8
11) #9 AND #10

**CENTRAL**

1) MeSH descriptor: [Patellofemoral Pain Syndrome] explode all trees
2) MeSH descriptor: [Chondromalacia Patellae] explode all trees
3) patell*:ti,ab OR femoropatell*:ti,ab OR retropatell*:ti,ab OR "anterior knee*":ti,ab OR peripatell*:ti,ab OR "kneecap*":ti,ab
4) pain*:ti,ab OR sore*:ti,ab OR discomfort*:ti,ab OR arthralgia*:ti,ab OR dysfunction*:ti,ab OR injur*:ti,ab OR syndr*:ti,ab OR chondromalac*:ti,ab OR chondropath*:ti,ab
5) MeSH descriptor: [Clinical Study] explode all trees OR MeSH descriptor: [Comparative Study] explode all trees OR MeSH descriptor: [Multicenter Study] explode all trees OR MeSH descriptor: [Cohort Studies] explode all trees
6) "randomised controlled trial*":ti,ab  OR "randomised clinical trial*":ti,ab  OR "randomized controlled trial*":ti,ab  OR "randomized clinical trial*":ti,ab OR "controlled trial*":ti,ab OR "controlled study*":ti,ab OR "clinical trial*":ti,ab OR "quasi-randomised":ti,ab  OR "quasi-randomized":ti,ab OR "semi-randomised":ti,ab OR "semi-randomized":ti,ab OR "observational stud*":ti,ab OR cohort*:ti,ab OR longitudinal*:ti,ab OR "prospective":ti,ab OR "long term follow-up":ti,ab OR "long-term follow-up":ti,ab OR "prognostic":ti,ab
7) # 1 OR #2
8) #3 AND #4
9) #5 OR #6
10) #7 OR #8
11) #9 AND #10

**Web of Science**

1) TS=(patell* OR femoropatell* OR femoro-patell* OR retropatell* OR retro-patell* OR "anterior knee*" OR peripatell* or peri-patell* OR "kneecap" OR patellofemoral OR patello-femoral OR "lateral compression syndrome*" OR "lateral facet syndrome*" OR "lateral pressure syndrome*" OR "lateral hyperpressure syndrome*" OR "odd facet syndrome*")
2) TS=(pain* OR sore* OR discomfort* OR arthralgia* OR dysfunction* OR injur* OR syndr* OR chondromalac* OR chondropath*)
3) TS=("randomised controlled trial*"OR "randomised clinical trial*"  OR "randomized controlled trial*" OR "randomized clinical trial*" OR "controlled trial*" OR "controlled study*" OR "clinical trial*" OR "quasi-randomised" OR "quasi-randomized" OR "semi-randomised" OR "semi-randomized" OR "observational stud*" OR cohort* OR longitudinal* OR "prospective" OR "long term follow-up" OR "long-term follow-up" OR "prognostic")
4) #1 AND #2
5) #3 AND #4

**Supplementary Appendix 2. Overview of extracted outcome measures**

| **Outcomes investigated, n (% of total studies)** | |
| --- | --- |
| **Pain outcome measures** | |
| Visual Analogue Scale (VAS) | 20 (47.6) |
| Presence of persistent symptoms | 10 (23.8) |
| Numeric Pain Rating Scale (NPRS) | 8 (19) |
| Pain intensity not specified | 4 (9.5) |
| Recurrence of pain | 1 (2.4) |
| **Improvement perception** | |
| Global Rating of Change (GROC) | 11 (26.2) |
| Patient satisfaction/perceived recovery | 11 (26.2) |
| Post operative grading (success, good, fair, poor etc) | 2 (4.8) |
| **Symptom, function and participation** | |
| Anterior Knee Pain Scale (AKPS)/Kujala | 17 (40.5) |
| Participation in sports/activities | 8 (19) |
| Knee injury and Osteoarthritis Outcome Score (KOOS) | 7 (16.7) |
| Functional Index Questionnaire (FIQ) | 4 (9.5) |
| Physical Activity Scale (PAS) | 2 (4.8) |
| Western Ontario and McMaster Universities Osteoarthritis Index (WOMAC) | 2 (4.8) |
| Lysholm Knee Scoring Scale | 2 (4.8) |
| Cincinnati Rating System (CRS) | 2 (4.8) |
| Lower Extremity Functional Scale (LEFS) | 1 (2.4) |
| Tegner activity scale | 1 (2.4) |
| Activities of Daily Living Scale (ADLS) | 1 (2.4) |
| VISA (Victorian Institute of Sport Assessment) score | 1 (2.4) |
| Feller’s Patella Score | 1 (2.4) |
| Non-specified function-reported outcomes | 2 (4.8) |
| **Psychosocial measures** | |
| Quality of Life (QoL) | 5 (11.9) |
| Hospital Anxiety and Depression (HAD) scale | 1 (2.4) |
| Tampa Scale of Kinesiophobia (TSK) | 1 (2.4) |
| Knee self-efficacy (KSES) | 1 (2.4) |
| **Physical/ clinical tests** | |
| Hip and/or knee strength | 6 (14.3) |
| Functional tests (single leg hops, step-down, unilateral squat etc) | 6 (14.3) |
| MRI/arthroscopic/ radiographic results/features | 3 (7.1) |
| Clinical tests (palpation, etc) | 2 (4.8) |
| Atrophy | 1 (2.4) |
| Patellar pressure | 1 (2.4) |
| Electromyography (EMG) analysis | 1 (2.4) |
| **Health-care utilization** | |
| Consumption of NSAIDs (nonsteroidal anti-inflammatory drugs)/medication | 7 (16.7) |
| Attended physiotherapist/General Practitioner consults | 2 (4.8) |

**Supplementary Appendix 3. GRADE ratings and justifications**

| **Outcomes** | **No. of studies (n)** | **Effect size (95% CI)** | **I²** | **Risk of bias** | **Inconsistency** | **Indirectness** | **Imprecision** | **Publication bias** | **Certainty (GRADE)** | **Reasons for downgrading** |
| --- | --- | --- | --- | --- | --- | --- | --- | --- | --- | --- |
| **Pain-related outcomes 12-month follow-up** |  |  |  |  |  |  |  |  |  |  |
| Worst pain | 7 (857) | SMD 1.36 (0.85 to 1.86) | 95.09% | Not serious | Serious | Not serious | Not serious | ? | ⊕⊕⊕⊝ MODERATE | Due to inconsistency (extreme heterogeneity: I2>90%) |
| Pain during activity | 6 (406) | SMD 1.36 (0.61 to 2.11) | 96.14% | Serious | Serious | Not serious | Serious | ? | ⊕⊕⊝⊝ LOW | Due to risk of bias, inconsistency (extreme heterogeneity: I2>90%), imprecision (wide CI) |
| Usual pain | 8 (616) | SMD 2.44 (−0.23 to 5.12) | 99.81% | Serious | Serious | Not serious | Serious | ? | ⊕⊕⊝⊝ LOW | Due to risk of bias, inconsistency (extreme heterogeneity: I2>90%), imprecision (wide/crosses null CI) |
| Resting pain | 5 (584) | SMD 0.91 (0.75 to 1.08) | 34.14% | Serious | Not serious | Not serious | Not serious | ? | ⊕⊕⊕⊝ MODERATE | Due to risk of bias |
| **24-month follow-up** |  |  |  |  |  |  |  |  |  |  |
| Usual pain | 3 (268) | SMD 4.44 (-3.05 to 11.92) | 99.86% | Serious | Serious | Not serious | Serious | ? | ⊕⊝⊝⊝ VERY LOW | Due to risk of bias, inconsistency (extreme heterogeneity: I2>90%), imprecision (wide/crosses null CI, small sample size (<300)) |
| **≥60-month follow-up** |  |  |  |  |  |  |  |  |  |  |
| Worst pain | 3 (157) | SMD 1.06 (0.50 to 1.62) | 82.17% | Serious | Serious | Not serious | Serious | ? | ⊕⊝⊝⊝ VERY LOW | Due to risk of bias, inconsistency (high heterogeneity: I2>75%), imprecision (small sample size: <300) |
| Pain during activity | 3 (702) | SMD 0.92 (0.76 to 1.07) | 54.12% | Very serious | Not serious | Not serious | Not serious | ? | ⊕⊕⊝⊝ LOW | Due to risk of bias |
| Usual pain | 3 (156) | SMD 1.11 (0.88 to 1.34) | 0% | Serious | Not serious | Not serious | Serious | ? | ⊕⊕⊝⊝ LOW | Due to risk of bias, imprecision: small sample size (<300) |
| **Function-related self-reported outcomes 12-month follow-up** |  |  |  |  |  |  |  |  |  |  |
| Physical function (AKPS) | 11 (1,015) | MD 14.60 (11.60 to 17.61) | 93.41% | Serious | Serious | Not serious | Not serious | ? | ⊕⊕⊝⊝ LOW | Due to risk of bias, inconsistency (extreme heterogeneity: I2>90%) |
| Physical function (FIQ) | 3 (509) | MD 3.33 (2.46 to 4.20) | 90.96% | Serious | Serious | Not serious | Not serious | ? | ⊕⊕⊝⊝ LOW | Due to risk of bias, inconsistency (extreme heterogeneity: I2>90%) |
| Physical function (WOMAC) | 2 (114) | MD –7.73 (- 10.36 to – 5.10) | 0% | Very serious | Not serious | Not serious | Serious | ? | ⊕⊕⊝⊝ LOW | Due to risk of bias, imprecision (small sample size: <300) |
| **≥60-month follow-up** |  |  |  |  |  |  |  |  |  |  |
| Physical function (AKPS and Lisholm) | 4 (195) | SMD -1.75 (-3.07 to -0.44) | 97.07% | Serious | Serious | Not serious | Serious | ? | ⊕⊝⊝⊝ VERY LOW | Due to risk of bias, inconsistency (extreme heterogeneity: I^2^>90%), imprecision (wide CI, small sample size: <300) |
| **Secondary outcomes  12-month follow-up** |  |  |  |  |  |  |  |  |  |  |
| KOOS Pain | 3 (192) | MD 10.48 (6.93 to 14.02) | 67.81% | Not serious | Not serious | Not serious | Serious | ? | ⊕⊕⊕⊝ MODERATE | Due to imprecision (wide CI and small sample size: <300) |
| KOOS Symptoms | 3 (192) | MD 4.71 (2.12 to 7.30) | 57.58 % | Not serious | Not serious | Not serious | Serious | ? | ⊕⊕⊕⊝ MODERATE | Due to imprecision (wide CI and small sample size: <300) |
| KOOS ADL | 3 (192) | MD 6.78 (2.52 to 11.08) | 78.14 % | Not serious | Serious | Not serious | Serious | ? | ⊕⊕⊝⊝ LOW | Due to inconsistency (high heterogeneity: I²>75%) and imprecision (wide CI and small sample size: <300) |
| KOOS Sports | 3 (192) | MD 13.31 (3.86 to 22.75) | 90.54 % | Not serious | Serious | Not serious | Serious | ? | ⊕⊕⊝⊝ LOW | Due to inconsistency (extreme heterogeneity: I^2^> 90%), and imprecision (wide CI and mall sample size: <300) |
| KOOS QoL | 3 (192) | MD 7.60 (5.94 to 9.25) | 0% | Not serious | Not serious | Not serious | Serious | ? | ⊕⊕⊕⊝ MODERATE | Due to imprecision (small sample size: <300) |
| GROC | 8 (1,078) | 0.54 (0.38 to 0.68) | 93.6% | Not serious | Serious | Not serious | Not serious | ? | ⊕⊕⊕⊝ MODERATE | Due to inconsistency (extreme heterogeneity: I^2^> 93%) |
| **Secondary outcomes  ≥60-month follow-up** |  |  |  |  |  |  |  |  |  |  |
| GROC | 3 (160) | 0.54 (0.43 to 0.65) | 52% | Serious | Not serious | Not serious | Serious | ? | ⊕⊕⊝⊝ LOW | Due to risk of bias, imprecision (small sample size: <300) |

**Supplementary Appendix 4. Meta-analysis of all subscales of Knee Injury and Osteoarthritis Outcome Score (KOOS) from baseline to 12 months follow-up.**

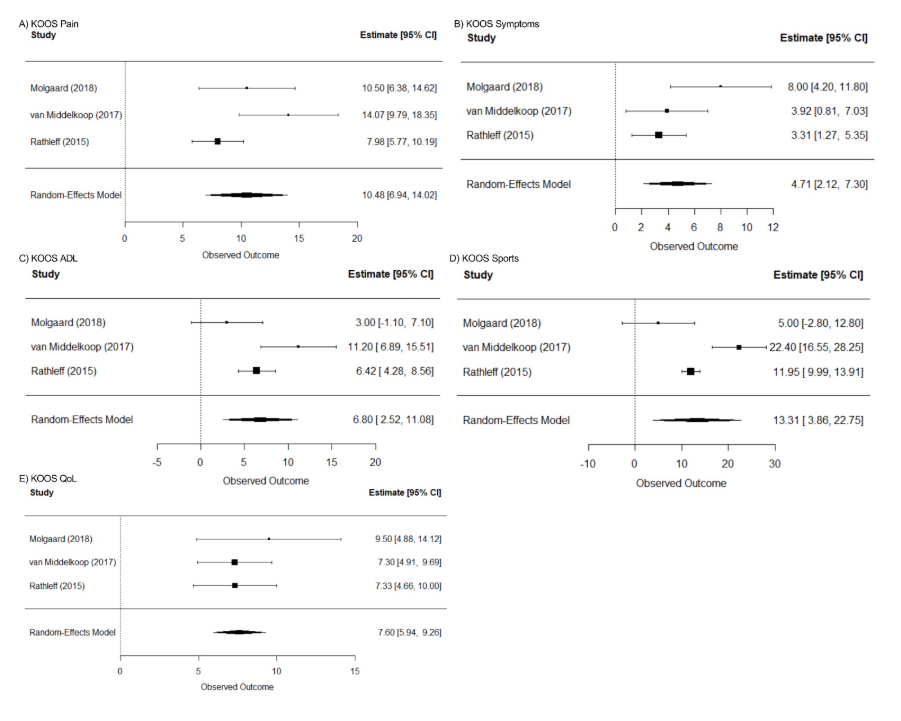

**Supplementary Appendix 5. Meta-analysis of improvement perception measured using Global Rating of Change (GROC), from baseline to 12 months and 60+ months follow-up.**

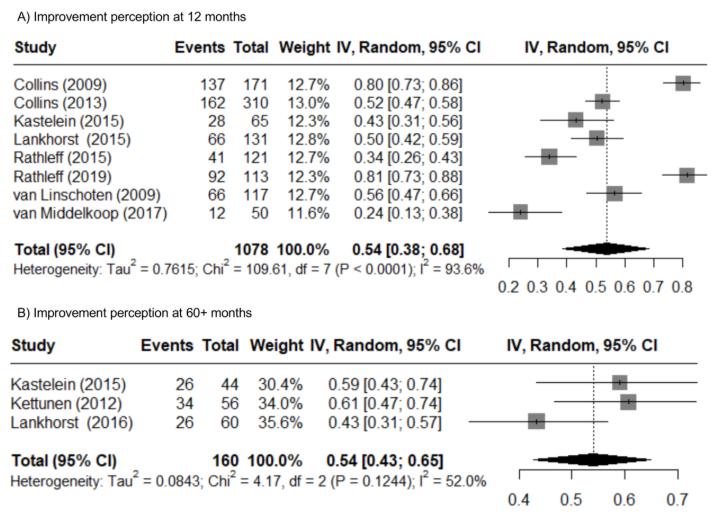
